# Why Depression Matters More for Dementia in Females: Global Population Evidence

**DOI:** 10.64898/2026.07.11.26357797

**Authors:** Wenpeng You

**Author notes:** Corresponding Author: Wenpeng You, PhD, MSc, BSc E.

## Abstract

Depression and dementia frequently co-occur, yet sex-specific population-level associations remain unclear. This ecological study examined cross-national relationships between sex-specific depressive disorder incidence and dementia incidence using Institute for Health Metrics and Evaluation data. Analyses included sex-stratified scatterplots, Pearson and Spearman correlations, principal component analysis, partial correlations adjusting for macro-structural factors, and sex-specific multiple linear regression. Visual analyses suggested positive associations in both sexes, but patterns were stronger and more structured among females. Female depressive disorder incidence correlated with dementia incidence in females and males, clustered with structural-development indicators, and remained associated after adjustment. Male depressive disorder incidence showed no significant associations. Overall, depressive disorder incidence was independently associated with dementia incidence among females but not males, supporting a sex-differentiated population-level mental health–dementia relationship with implications for global women’s health and ageing. These findings inform sex-sensitive surveillance, prevention, and policy frameworks in rapidly ageing societies worldwide, particularly within resource-constrained transitional contexts.

## Background

Dementia is a leading cause of disability and dependency worldwide, with incidence continuing to rise alongside population ageing and extended life expectancy (WHO, 2021, 2025). Although biological ageing is a fundamental driver of dementia, substantial cross-national variation in dementia incidence indicates that structural, social, and health-related factors also play an important role (Giannoni-Luza et al., 2025; Rodriguez et al., 2008). Understanding how these factors operate differently across population subgroups, particularly by sex, is essential for refining dementia epidemiology and informing prevention strategies (Huque et al., 2023; Rocca et al., 2014).

Depressive disorders have long been implicated in dementia risk, yet their role remains contested (Ownby et al., 2006). At the individual level, depression has been associated with accelerated cognitive decline and increased dementia risk (Ownby et al., 2006; Saczynski et al., 2010), but findings are inconsistent due to heterogeneity in study design, timing of exposure, follow-up duration, and confounder adjustment (Byers & Yaffe, 2011; Ownby et al., 2006). Depression has been interpreted as a prodromal manifestation of neurodegeneration, a comorbid condition arising from shared biological or vascular mechanisms, or a marker of broader psychosocial vulnerability (Barnes et al., 2006; Prince et al., 2013; Singh-Manoux et al., 2017). These complexities limit causal inference and highlight the value of complementary population-level analyses examining how depressive disorder incidence and dementia incidence co-vary across countries.

Sex differences represent a critical but underexplored dimension of the depression–dementia relationship (Fuhrer et al., 2003; Gong et al., 2023; Nebel et al., 2018). Globally, females experience higher incidence of depressive disorders and a greater burden of dementia than males, even after accounting for longer life expectancy (Fuhrer et al., 2003). Biological mechanisms such as hormonal and immune differences have been proposed (Nebel et al., 2018). However, social and structural factors also vary systematically by sex and across societies (Rocca et al., 2014; Solar & Irwin, 2010). Most population level studies nevertheless pool sexes or treat sex as a covariate, rather than directly examining sex differentiated associations (Ferretti et al., 2018; Nebel et al., 2018).

At the macro-structural level, dementia incidence is strongly associated with indicators of population ageing and socioeconomic development, including life expectancy at older ages, economic affluence, urbanisation, and reduced exposure to premature mortality (Li et al., 2022; Nichols et al., 2022; You & Henneberg, 2026). Depressive disorder incidence also varies widely across countries and is shaped by healthcare access, diagnostic capacity, urban living environments, and cultural norms surrounding mental health reporting (Ferrari et al., 2013; Peen et al., 2010). Consequently, observed associations between depression and dementia may reflect shared structural contexts rather than direct mental health pathways (Singh-Manoux et al., 2017).

Emerging evidence suggests that depressive disorder incidence may be embedded differently within these macro-structural contexts for females and males (King et al., 2020; Van de Velde et al., 2010). Female mental health outcomes appear more closely aligned with socioeconomic development, longevity gains, and demographic transition (Levinson et al., 2009; Riecher-Rössler, 2017), whereas male depressive disorder incidence often shows weaker or inconsistent associations with these structural indicators (Levinson et al., 2009). This divergence raises the possibility that depressive disorder incidence functions as a population-level vulnerability marker for dementia in females but not in males (Fuhrer et al., 2003; Livingston et al., 2024).

A direct sex-comparative analysis therefore offers an opportunity to clarify whether depressive disorder operates as a shared correlate of dementia or as a sex-specific population-level indicator (Diez Roux, 2004; Nebel et al., 2018). By explicitly comparing depressive disorder–dementia associations in females and males, it becomes possible to disentangle structural co-variation from independent associations and assess whether mental health contributes differentially to dementia burden by sex (Fuhrer et al., 2003).

Few studies have systematically undertaken such sex-stratified population-level comparisons using multiple complementary analytical approaches or examined whether observed differences persist after adjustment for key macro-structural covariates. The present study addresses these gaps through a global ecological analysis comparing depressive disorder–dementia relationships in females and males. Using visualisation, correlation analyses, principal component analysis, partial correlations, and multivariable regression, this study evaluates whether depressive disorder incidence contributes differential explanatory value for dementia incidence by sex and whether it functions as a sex-differentiated population-level vulnerability indicator rather than a uniform mental health correlate.

## Materials and Methods

### Data sources

Sex-stratified country-level incidence data for dementia and depressive disorders were obtained from the Global Burden of Disease (GBD) Study curated by the Institute for Health Metrics and Evaluation (IHME), University of Washington, USA (IHME, 2023; Karlsson et al., 2014). Dementia incidence estimates for 2023, including Alzheimer’s disease and other dementias, were derived separately for females and males using standard GBD modelling approaches that synthesise population-based cohort studies, health surveys, hospital and outpatient records, administrative claims, and cause-of-death data. Case definitions followed International Classification of Diseases (ICD) criteria for Alzheimer’s disease and related dementias (IHME, 2023; Karlsson et al., 2014).

Depressive disorder incidence rates (new cases per 100,000 population in 2018) were obtained from GBD depressive disorder estimates based on population registries, surveys, vital registration systems, and hospital data, synthesised using established GBD statistical methods (IHME, 2023; Karlsson et al., 2014).

Macro-structural indicators were compiled from international sources. Economic affluence was measured using gross domestic product (GDP) per capita adjusted for purchasing power parity (PPP; constant 2020 US dollars) (You & Henneberg, 2017), and urbanisation was defined as the percentage of the population residing in urban areas (Bray et al., 2024; Li et al., 2023); both were obtained from the World Bank World Development Indicators. Life expectancy at age 60 (e₆₀; years in 2020) for females and males was retrieved from the World Health Organization (WHO) Global Health Observatory (Nichols et al., 2022). Henneberg Index, the Biological State Index (I_bs_), reflecting population-level exposure to natural selection through mortality and fertility rates, derived from previously published estimates (You & Henneberg, 2026).

Countries were classified using multiple international frameworks to enable stratified analyses, including World Bank income categories, United Nations development status (developed vs developing), and the six WHO regions. Additional geopolitical and cultural groupings (e.g., APEC, Arab World, European Union, OECD, Latin America and the Caribbean, SADC, SCO, and English-speaking countries) were included to capture broader contextual variation. Analyses were based on data from up to 204 countries, with sample sizes varying according to data completeness.

All data used in this study were secondary data published by international organisations. As no individual-level or identifiable data were involved, informed consent was not required.

### Study design and variable specification

This study employed a sex-stratified ecological, cross-national design. All analyses were conducted separately for females and males to enable direct comparison of population-level associations. Depressive disorder incidence was specified as the primary explanatory variable and dementia incidence as the outcome.

This specification was guided by epidemiological and life-course considerations: depressive disorders are typically diagnosed earlier in adulthood, whereas dementia is predominantly a late-life condition. Sex stratification therefore allows assessment of whether population-level depressive disorder–dementia associations differ between females and males, rather than assuming a uniform pathway.

Except for the Henneberg Index, which was derived from a single dataset, all variables showed consistently strong country level correlations over the period 2009 to 2023, indicating stable national patterns over time. This temporal stability supports interpreting the exposures as reflecting enduring structural, socioeconomic, mental health, and longevity contexts rather than short-term fluctuations. Accordingly, the specific year of measurement is unlikely to materially affect the ecological associations examined in this study.

### Data preparation

Data from multiple international repositories were harmonised to construct a comprehensive country-level dataset. Incidence measures and macro-structural indicators (GDP PPP, urbanisation, Henneberg Index, and e*_60_*) were aligned by country and expressed using standardised units.

Countries with missing data on any study variable were excluded using listwise deletion to ensure consistent analytic samples across correlation, principal component, partial correlation, and regression analyses.

Given the skewed distributions typical of cross-national incidence and development indicators, all incidence and macro-structural variables were log₁₀-transformed prior to analysis to reduce skewness, stabilise variance, and limit the influence of extreme values at higher incidence levels.

Multicollinearity among explanatory variables was assessed separately in female and male regression models using tolerance and variance inflation factor (VIF) statistics. All models met conventional thresholds (tolerance ≥ 0.10; VIF ≤ 10), indicating no problematic multicollinearity.

### Statistical analysis

Scatterplots were first examined to assess distributions and potential non-linearity. Pearson’s correlation coefficients and Spearman’s rank correlations were used to examine associations across all countries and within predefined country groupings. Differences in correlation strength across income levels and development status were tested using Fisher’s r-to-z transformations.

Principal component analysis (PCA) was conducted separately for females and males to assess whether depressive disorder incidence clustered with indicators of population ageing and socioeconomic development (GDP PPP, urbanisation, Henneberg Index, and e*_60_*). Sampling adequacy was evaluated using the Kaiser–Meyer–Olkin statistic, and Bartlett’s test of sphericity was used to assess factorability. Components with eigenvalues greater than one were retained.

Partial correlation analyses were performed to assess whether depressive disorder–dementia associations persisted after sequential adjustment for macro-structural covariates, including sex-specific life expectancy at age 60. Multiple linear regression models were then estimated separately for females and males using the ENTER method to compare explanatory models with and without depressive disorder incidence. Standardised regression coefficients (β), adjusted R² values, and F-statistics were reported.

Except for the scatter plots generated in Microsoft Excel® 2016, all variables were log₁₀-transformed prior to analysis. All statistical analyses were conducted using IBM SPSS Statistics, version 31 (IBM Corp., 2023). Findings are interpreted at the population level and do not imply causality or individual-level risk.

## Results

### Visual sex-specific associations

Figure 1 presents sex-specific scatterplots illustrating the population-level association between depressive disorder incidence and dementia incidence across countries. In both sexes, higher depressive disorder incidence was generally associated with higher dementia incidence; however, the strength and explanatory value of these associations differed substantially by sex.

**Figure 1:**
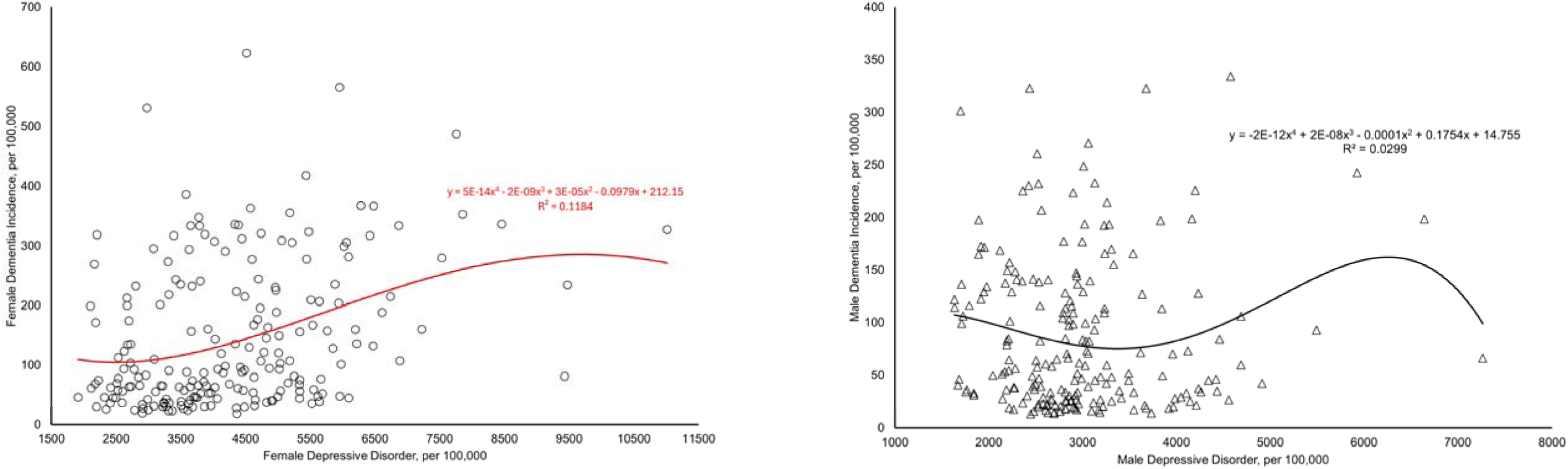
Sex-specific scatter plots of depressive disorder and dementia incidence. Data sources: Depressive incidence rate and dementia incidence rate (including Alzheimer’s disease and other dementias), for both sexes, were obtained from the Institute for Health Metrics and Evaluation (IHME), University of Washington. Incidence rates are reported as the number of new cases per 100,000 population. Quadratic regression lines are shown for illustrative purposes; regression equations and R² values are displayed within each panel.

Among females, the association was stronger and more structured, with a clear non-linear increase in dementia incidence at higher levels of depressive disorder incidence. The quadratic model explained a meaningful proportion of between-country variation in female dementia incidence (R² = 0.118), indicating a more coherent population-level relationship.

In contrast, the association among males was weak and highly dispersed, with considerable variability in dementia incidence across similar levels of depressive disorder incidence. The quadratic model demonstrated very limited explanatory power (R² = 0.030), suggesting that depressive disorder incidence accounted for little cross-national variation in male dementia incidence.

Overall, these visual patterns indicate a pronounced sex difference, with depressive disorder incidence more strongly associated with dementia incidence in females than in males at the population level. Quadratic regression lines are shown for descriptive purposes only and do not imply causality.

### Bivariate correlation Matrix

Bivariate correlation analyses revealed a marked sex difference in the population-level association between depressive disorder incidence and dementia incidence (Table 1). Female depressive disorder incidence was consistently and positively associated with dementia incidence in both females (Pearson’s r = 0.340; Spearman’s ρ = 0.341, both p < .001) and males (r = 0.383; ρ = 0.380, both p < .001). In contrast, male depressive disorder incidence showed no significant association with dementia incidence in either females or males across both correlation metrics.

**Table 1.**
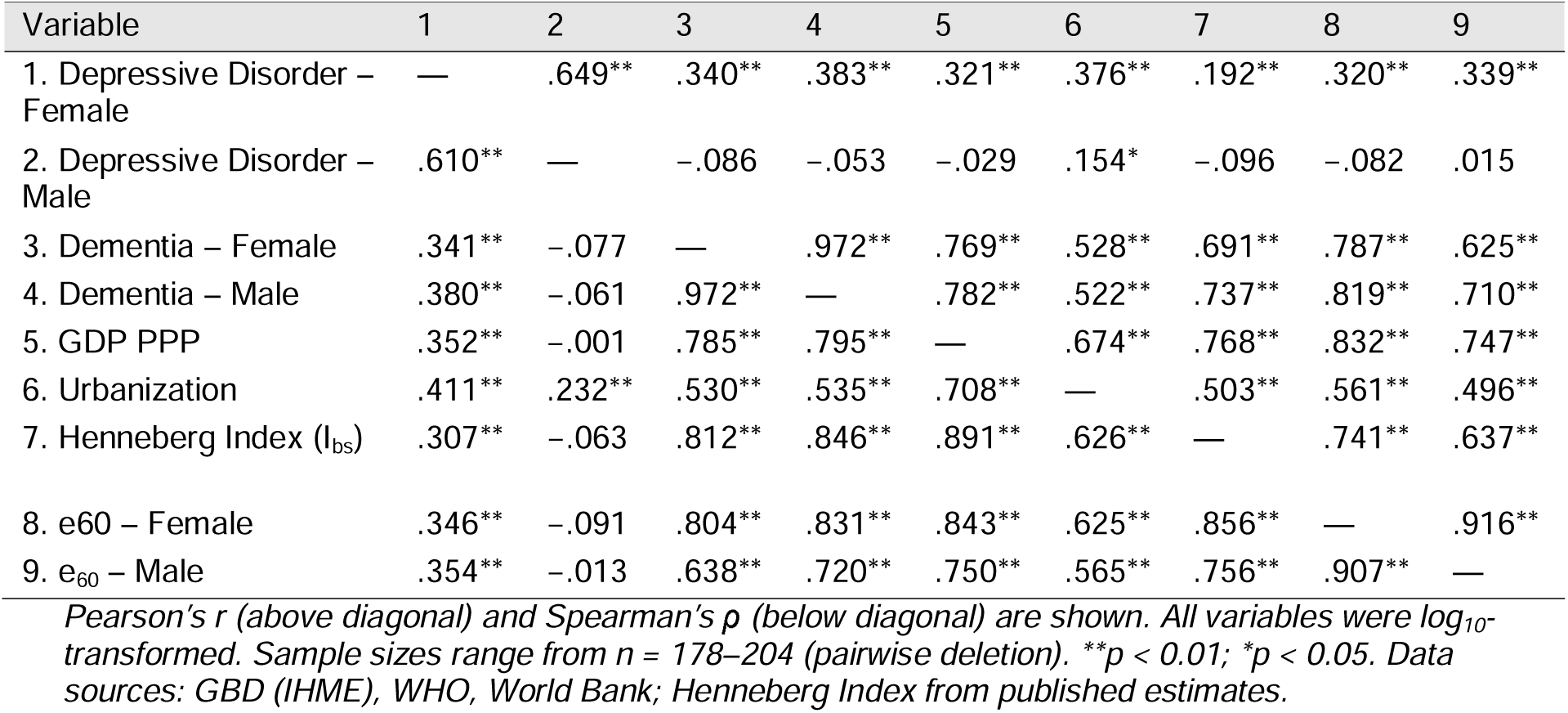
Bivariate correlations between depressive disorder incidence, dementia incidence, and key macro-level covariates Principal component structure of depressive disorder incidence and structural covariates.

| Variable | 1 | 2 | 3 | 4 | 5 | 6 | 7 | 8 | 9 |
| --- | --- | --- | --- | --- | --- | --- | --- | --- | --- |
| 1. Depressive Disorder – Female | — | .649** | .340** | .383** | .321** | .376** | .192** | .320** | .339** |
| 2. Depressive Disorder – Male | .610** | — | -.086 | -.053 | -.029 | .154* | -.096 | -.082 | .015 |
| 3. Dementia – Female | .341** | -.077 | — | .972** | .769** | .528** | .691** | .787** | .625** |
| 4. Dementia – Male | .380** | -.061 | .972** | — | .782** | .522** | .737** | .819** | .710** |
| 5. GDP PPP | .352** | -.001 | .785** | .795** | — | .674** | .768** | .832** | .747** |
| 6. Urbanization | .411** | .232** | .530** | .535** | .708** | — | .503** | .561** | .496** |
| 7. Henneberg Index ($I_{bs}$) | .307** | -.063 | .812** | .846** | .891** | .626** | — | .741** | .637** |
| 8. e60 – Female | .346** | -.091 | .804** | .831** | .843** | .625** | .856** | — | .916** |
| 9. e<sub>60</sub> – Male | .354** | -.013 | .638** | .720** | .750** | .565** | .756** | .907** | — |

*Pearson's $r$ (above diagonal) and Spearman's $\rho$ (below diagonal) are shown. All variables were $\log_{10}$ -transformed. Sample sizes range from $n = 178$ – $204$ (pairwise deletion). \*\* $p < 0.01$ ; \* $p < 0.05$ . Data sources: GBD (IHME), WHO, World Bank; Henneberg Index from published estimates.*

Dementia incidence in females and males was highly correlated (r = 0.972, p < .001), reflecting a shared cross-national dementia burden. However, this strong co-variation was not mirrored by male depressive disorder incidence, indicating a sex-specific divergence in the depression–dementia relationship. Formal comparison using Fisher’s r-to-z transformation confirmed that the depressive disorder–dementia association was significantly stronger in females than in males for both Pearson’s (z = 4.08, p < .001) and Spearman’s correlations (z = 4.17, p < .001).

Dementia incidence in both sexes showed strong positive correlations with macro-level indicators of population ageing and development, including life expectancy at age 60, GDP PPP, and reduced selection opportunity (all r > 0.69, p < .001), with moderate associations observed for urbanisation. Female depressive disorder incidence exhibited similar positive correlations with these structural indicators, whereas male depressive disorder incidence demonstrated weak or null associations, apart from a modest correlation with urbanisation.

Sex-specific correlation patterns across global, economic, and regional country groupings are summarised in Supplementary Table S1. At the global level, a moderate and statistically significant positive association was consistently observed among females, whereas male associations remained weak or non-significant. Across income groups and development classifications, female correlations were broadly stable and tended to strengthen in lower-income contexts, while male correlations showed no consistent pattern. Substantial regional heterogeneity was evident, with stronger female associations in the Eastern Mediterranean and Western Pacific regions and largely null findings in Europe and South-East Asia. Overall, these results indicate a robust and predominantly sex-differentiated ecological association between depressive disorder incidence and dementia incidence across diverse global contexts.

PCA was used descriptively to assess whether depressive disorder incidence clustered with macro-structural indicators of population ageing and socioeconomic development, and whether this structure differed by sex (Table 2). In females, a single component explained 64.1% of the variance (KMO = 0.778; Bartlett’s test p < .001) and was dominated by GDP PPP, urbanisation, reduced selection opportunity, and female life expectancy at age 60. Female depressive disorder incidence loaded moderately on this component (loading = 0.463) with low communality (0.214), indicating partial structural embedding. In males, a two-component solution explained 79.7% of the variance (KMO = 0.746; Bartlett’s test p < .001). Male depressive disorder incidence defined a distinct second component (loading = 0.975; communality = 0.951), largely independent of structural indicators. These patterns align with subsequent analyses, consistent with a sex-differentiated population-level vulnerability interpretation rather than a uniform mental health correlate.

**Table 2.**
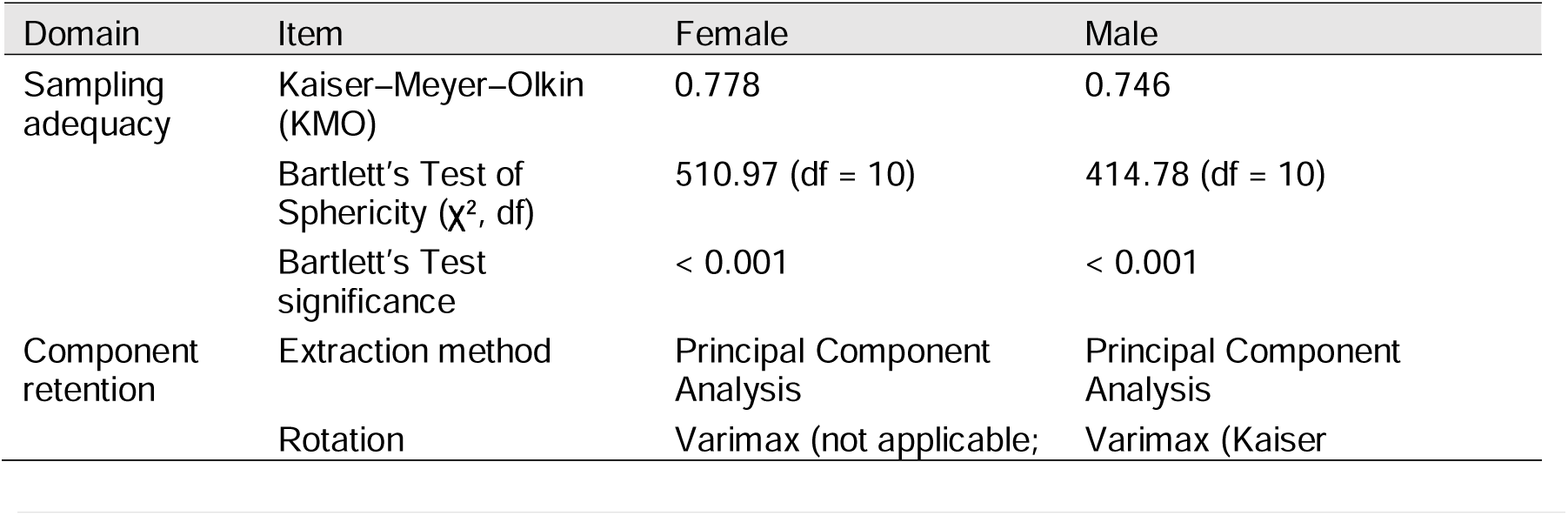
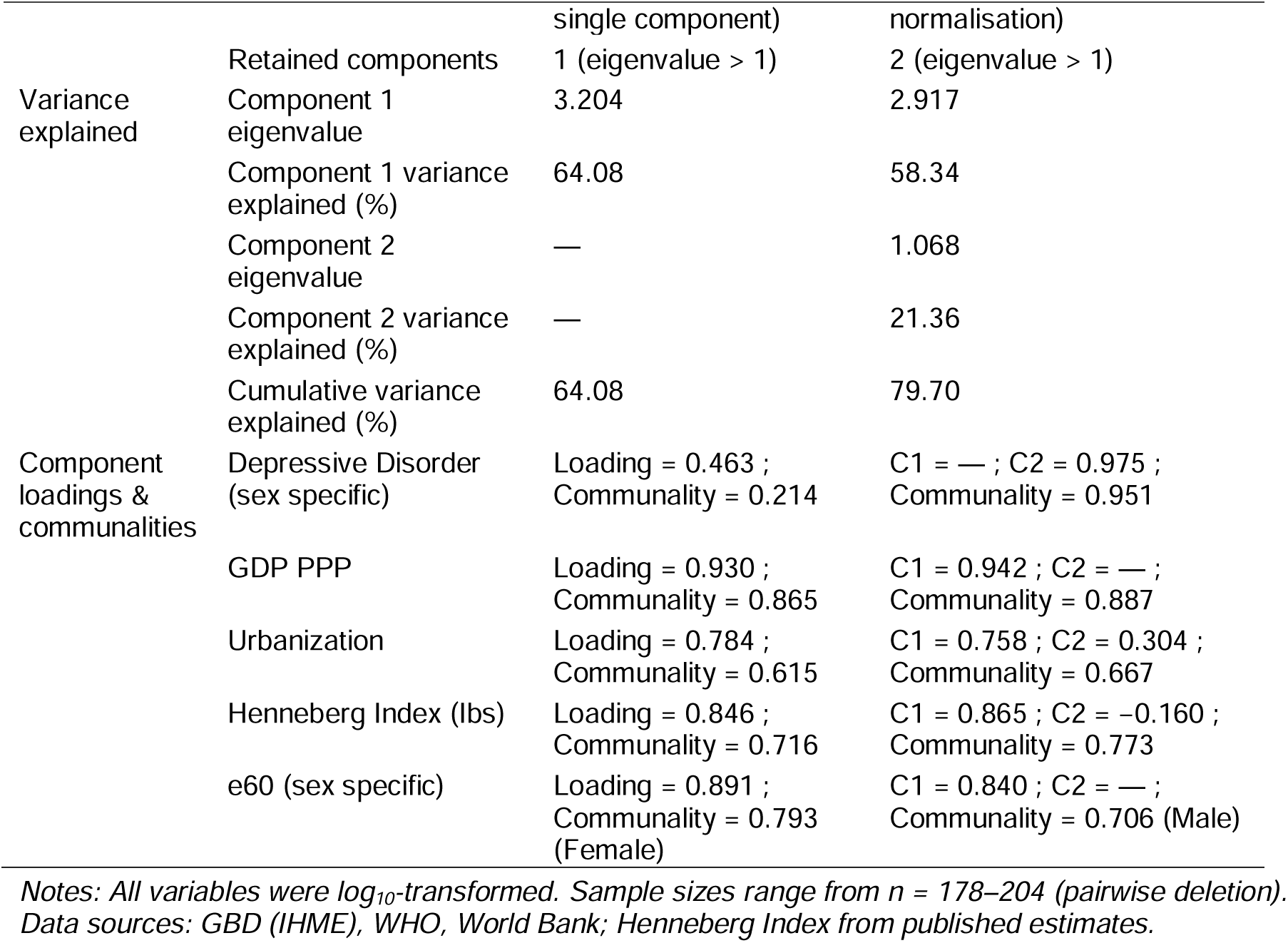
Principal component analysis of depression incidence and structural covariates by sex.

### Partial correlations between depressive disorder incidence and dementia incidence

Partial correlation analyses demonstrated a clear sex difference in the independence of the depressive disorder–dementia association (Table 3). At the zero-order level, depressive disorder incidence was moderately and positively associated with dementia incidence among females (r = 0.319, p < .001), whereas no association was observed among males (r = −0.044, p = .561). This female–male difference was statistically significant (Fisher’s z = 3.46, p < .001).

**Table 3.**
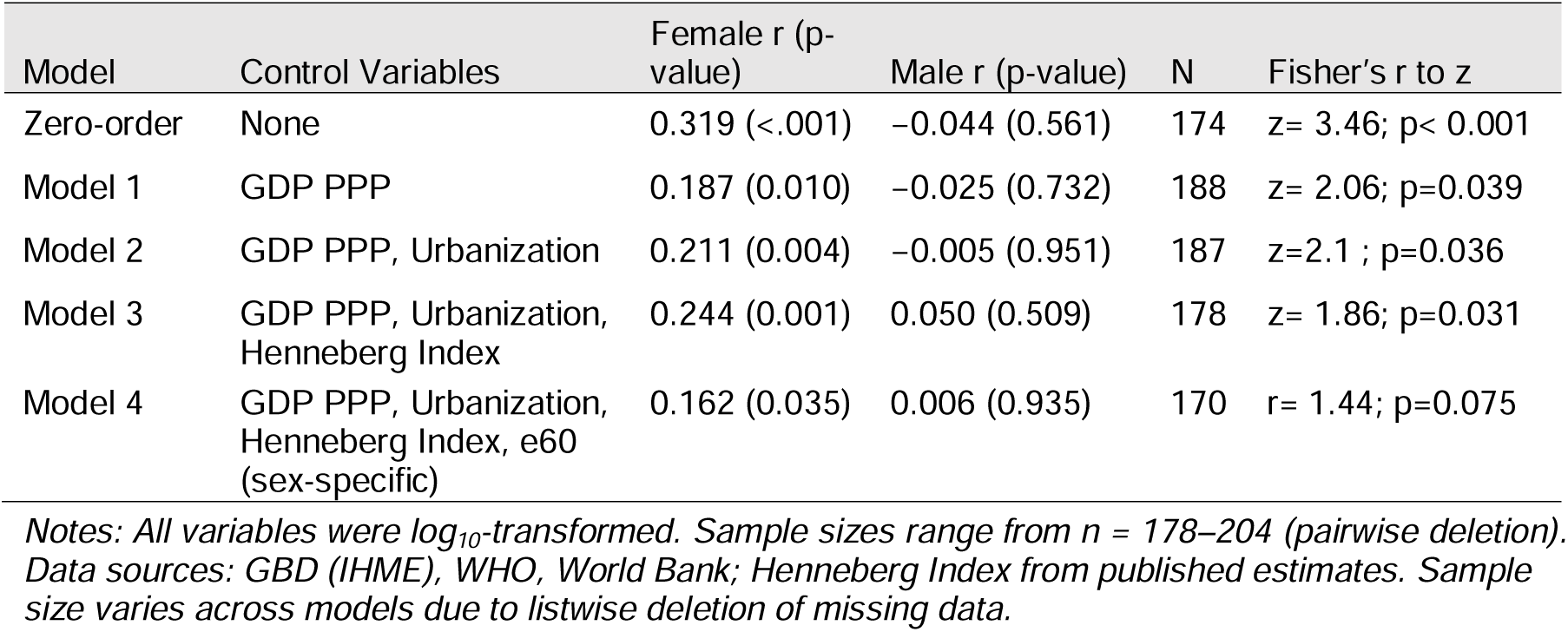
Partial correlations between depressive disorder and dementia incidence by sex, with confounders held statistically constant.

| Model | Control Variables | Female r (p-value) | Male r (p-value) | N | Fisher's r to z |
| --- | --- | --- | --- | --- | --- |
| Zero-order | None | 0.319 (<.001) | -0.044 (0.561) | 174 | z= 3.46; p< 0.001 |
| Model 1 | GDP PPP | 0.187 (0.010) | -0.025 (0.732) | 188 | z= 2.06; p=0.039 |
| Model 2 | GDP PPP, Urbanization | 0.211 (0.004) | -0.005 (0.951) | 187 | z=2.1 ; p=0.036 |
| Model 3 | GDP PPP, Urbanization, Henneberg Index | 0.244 (0.001) | 0.050 (0.509) | 178 | z= 1.86; p=0.031 |
| Model 4 | GDP PPP, Urbanization, Henneberg Index, e60 (sex-specific) | 0.162 (0.035) | 0.006 (0.935) | 170 | r= 1.44; p=0.075 |

*Notes: All variables were $\log_{10}$ -transformed. Sample sizes range from $n = 178$ – $204$ (pairwise deletion). Data sources: GBD (IHME), WHO, World Bank; Henneberg Index from published estimates. Sample size varies across models due to listwise deletion of missing data.*

After adjustment for economic affluence (GDP PPP), the association remained significant in females (r = 0.187, p = .010) but not in males (r = −0.025, p = .732), with the female association remaining significantly stronger. Further adjustment for urbanisation and reduced selection opportunity produced stable and significant associations in females (r = 0.211–0.244, p ≤ .004), while associations in males remained consistently null. Fisher’s r-to-z tests continued to indicate stronger associations in females across these models.

Following additional adjustment for sex-specific life expectancy at age 60, the association in females was attenuated but remained statistically significant (r = 0.162, p = .035), whereas the male association remained non-significant (r = 0.006, p = .935). The corresponding sex difference was attenuated and no longer statistically significant (z = 1.44, p = .075), suggesting that longevity partially accounts for, but does not fully explain, the stronger female association.

Overall, these findings indicate that depressive disorder incidence is independently associated with dementia incidence in females across multiple structural adjustments, whereas no independent association is evident in males. The persistence of the female association, alongside an attenuated and non-significant sex difference after longevity adjustment, supports a predominantly sex-differentiated population-level link that is partly mediated by ageing-related factors.

### Multiple linear regression analysis of depressive disorder incidence and dementia incidence: sex comparison

Multiple linear regression analyses demonstrated a clear sex difference in the contribution of depressive disorder incidence to dementia incidence after adjustment for macro-structural covariates (Table 4).

**Table 4.** Multiple linear regression (enter) predicting dementia incidence by sex.

| Predictor | Female Model 1 $\beta$<br>(p) | Female Model 2 $\beta$<br>(p) | Male Model 1 $\beta$<br>(p) | Male Model 2 $\beta$<br>(p) |
| --- | --- | --- | --- | --- |
| GDP PPP | 0.265 (0.008) | 0.280 (0.005) | 0.350 (<0.001) | 0.349 (<0.001) |
| Urbanization | 0.037 (0.537) | -0.001 (0.991) | 0.003 (0.963) | 0.005 (0.940) |
| Henneberg Index | 0.163 (0.027) | 0.175 (0.018) | 0.332 (<0.001) | 0.331 (<0.001) |
| e60 (sex-specific) | 0.419 (<0.001) | 0.392 (<0.001) | 0.225 (<0.001) | 0.226 (<0.001) |
| Depressive Disorder<br>(sex-specific) | — | 0.088 (0.077) | — | -0.005 (0.907) |
| Model $R^2$ / Adj. $R^2$ | 0.664 / 0.656 | 0.670 / 0.660 | 0.678 / 0.670 | 0.678 / 0.668 |
| F (df) | 83.32 (4,169) | 68.15 (5,168) | 88.77 (4,169) | 70.61 (5,168) |
| Model p-value | <0.001 | <0.001 | <0.001 | <0.001 |

*Notes: Models were estimated using the ENTER method with listwise deletion (N = 174). All variables were $\log_{10}$ -transformed. Sample sizes range from n = 178–204 (pairwise deletion). Data sources: GBD (IHME), WHO, World Bank; Henneberg Index from published estimates.*

Among females, the structural covariate model explained 66.4% of the variance in dementia incidence (R² = 0.664, p < .001), with life expectancy at age 60 emerging as the strongest predictor, followed by GDP PPP and reduced selection opportunity. When female depressive disorder incidence was added to the model, explained variance increased modestly (R² = 0.670; ΔR² = 0.006), and depressive disorder incidence showed a positive, borderline independent association with female dementia incidence (β = 0.088, p = .077).

In contrast, among males, the structural covariate model explained a similar proportion of variance (R² = 0.678, p < .001), driven primarily by reduced selection opportunity, GDP PPP, and life expectancy at age 60. Inclusion of male depressive disorder incidence did not improve model fit (ΔR² ≈ 0), and depressive disorder incidence was not independently associated with male dementia incidence (β = −0.005, p = .907).

Comparison of sex-stratified models therefore indicates that depressive disorder incidence contributes additional explanatory value for dementia incidence in females but not in males, even after accounting for shared structural and longevity-related determinants. These findings reinforce the interpretation of depressive disorder incidence as a female-specific population-level correlate of dementia rather than a uniform predictor across sexes.

## Discussion

This study demonstrates that depressive disorder incidence operates differently within population-level dementia risk structures for females and males. Rather than constituting a uniform mental health correlate of dementia, depressive disorder incidence appears to function as a sex-specific population-level indicator, with relevance primarily for female dementia burden. This finding refines existing dementia epidemiology by showing that population-level mental health–dementia relationships are shaped by sex-specific structural embedding rather than reflecting a single shared pathway.

At the individual level, depression has long been linked to dementia risk, yet the nature of this relationship remains contested (Ownby et al., 2006; Saczynski et al., 2010). Prior cohort and registry studies have variably interpreted depression as a prodromal manifestation of neurodegeneration, a shared-risk condition arising from vascular or inflammatory mechanisms, or an independent psychosocial risk factor (Alexopoulos et al., 1997; Fuhrer et al., 2003; Richard et al., 2013). However, effect sizes are inconsistent and frequently attenuate after adjustment for ageing, socioeconomic position, and comorbidities (Saczynski et al., 2010; Stafford et al., 2022). Importantly, most previous studies have either pooled sexes or statistically adjusted for sex, limiting insight into whether depression-related dementia risk operates similarly for women and men (Cherbuin et al., 2015; Nebel et al., 2018; You, 2025b). By adopting a sex-comparative ecological framework, the present study extends this literature by demonstrating that depressive disorder incidence aligns with dementia burden in a fundamentally different way across sexes at the population level.

Multivariable regression analyses further supported this sex-specific pattern. After adjustment for macro-structural covariates, depressive disorder incidence contributed modest additional explanatory value to female dementia incidence (ΔR² = 0.006), with no corresponding contribution observed in males. While the independent contribution in the female regression model was modest, its relevance is supported by consistent sex-specific patterns observed across multiple analytical frameworks, which is central to inference in ecological studies.

The sex-differentiated association observed in this study suggests that depressive disorder incidence captures broader population-level vulnerabilities that are relevant to dementia burden beyond ageing alone (You et al., 2025). In women, depressive disorder incidence appears partially embedded within macro-structural contexts characterised by longevity gains, socioeconomic development, and reduced exposure to premature mortality (Christiansen et al., 2022; Levinson et al., 2009; Patel et al., 2018). This embedding likely reflects cumulative life-course exposures, including differential access to education, labour participation, caregiving roles, and social protection, which are known to vary systematically by sex across societies (Ben-Shlomo & Kuh, 2002; Kuh et al., 2003). At the population level, depressive disorder incidence among females may therefore act less as a discrete psychiatric signal and more as a marker of accumulated psychosocial and structural stress that becomes increasingly salient as survival into older age improves (Ben-Shlomo & Kuh, 2002; You, 2025a).

In contrast, the absence of a comparable association among males suggests that male depressive disorder incidence does not function as a stable population-level marker of dementia burden. This does not imply that depression is irrelevant to dementia risk in men at the individual level, but rather that male depressive disorder incidence is less structurally integrated within demographic and developmental processes that shape cross-national dementia patterns (Diez Roux, 2004; Fuhrer et al., 2003; Levinson et al., 2009). Gender norms, underdiagnosis, stigma, and differences in health-seeking behaviour may weaken the correspondence between recorded depressive disorder incidence and underlying psychosocial burden among men, particularly in low-resource settings (Addis & Mahalik, 2003; Courtenay, 2000). As a result, male depressive disorder incidence may be a less reliable ecological indicator of downstream cognitive health outcomes.

The global distribution of dementia further contextualises these findings. According to the World Health Organization, approximately 60% of people living with dementia currently reside in low- and middle-income countries, where populations are undergoing rapid demographic transition under persistent structural constraints (Prince et al., 2015; WHO, 2021). In many of these settings, women experience disproportionate cumulative disadvantage across the life course, including limited educational opportunities, economic dependency, unpaid caregiving responsibilities, and restricted access to mental health care (Mikkola, 2005; WHO, 2008). Within such contexts, female depressive disorder incidence may function as a population-level signal of compounded psychosocial and structural vulnerability that intersects with ageing processes to shape dementia burden (King et al., 2020; Krieger, 2024). The stronger female-specific associations observed in developing and structurally transitioning contexts are therefore consistent with broader global patterns of gendered health inequality (King et al., 2020; WHO, 2008).

These findings also contribute to ongoing debates regarding the interpretation of depression–dementia associations (Gong et al., 2023; Ownby et al., 2006). While individual-level studies have struggled to disentangle causality from shared risk and reverse pathways (Byers & Yaffe, 2011), population-level analyses offer complementary insight into how mental health conditions are embedded within structural environments (Diez Roux, 2004; Krieger, 2024). The present study suggests that, at the ecological level, depressive disorder incidence does not uniformly reflect dementia risk but instead interacts with sex-specific social and demographic structures. This distinction highlights the importance of separating individual-level mechanisms from population-level indicators when interpreting mental health–dementia relationships (Diez Roux, 2004).

From a methodological perspective, the study highlights the value of explicit sex comparison rather than statistical adjustment alone. Treating sex as a covariate assumes equivalence in underlying pathways, whereas direct comparison reveals meaningful divergence in how mental health conditions relate to dementia burden across populations (Clayton & Tannenbaum, 2016; Nebel et al., 2018). The findings caution against extrapolating pooled or sex-adjusted estimates to global prevention frameworks without considering sex-specific structural contexts (Heidari et al., 2016; Organization, 2019).

In summary, this study shows that depressive disorder incidence functions as a sex-differentiated population-level correlate of dementia, with primary relevance to female dementia burden. Rather than indicating a direct or universal causal pathway, this association appears to reflect the intersection of mental health, longevity, and cumulative structural disadvantage (Ben-Shlomo & Kuh, 2002; Krieger, 2024). Recognising these sex-specific patterns is essential for refining interpretation of global dementia epidemiology and for informing prevention frameworks that are sensitive to gendered life-course and structural contexts, particularly in regions where the majority of the world’s dementia burden resides. By explicitly comparing females and males rather than treating sex as a covariate, this study underscores the value of sex-stratified approaches for identifying population-level vulnerability markers in dementia research.

From a women’s health perspective, the findings suggest that depressive disorder incidence may act as a population-level signal of cumulative psychosocial and structural vulnerability among women as survival into older age improves. Across many settings, women experience lifelong gendered disadvantage, including lower educational attainment, economic dependency, unpaid caregiving responsibilities, and constrained access to mental health care (Mikkola, 2005; WHO, 2008). These conditions may both elevate depressive disorder incidence and shape cognitive ageing trajectories through sustained psychosocial stress, reduced cognitive reserve, and delayed access to preventive services [37, 38]. Integrating sex-sensitive mental health surveillance into dementia prevention strategies may help identify populations of women at heightened risk, particularly in low- and middle-income countries undergoing rapid demographic transition (King et al., 2020; Krieger, 2024).

### Strengths and limitations

This study has several notable strengths. It adopts an explicitly sex-comparative population-level design rather than treating sex as a covariate, enabling direct assessment of whether depressive disorder–dementia associations differ between females and males. This approach allowed identification of a female-specific pattern that would likely have been obscured in pooled analyses. Multiple complementary analytical methods were applied. These included visualisation, bivariate correlations, principal component analysis, partial correlations, and multivariable regression. Together, these approaches provided convergent evidence for the observed sex differences. Key macro structural and longevity related covariates were included. These comprised economic development, urbanisation, reduced selection opportunity, and sex specific e*_60_*. This approach allowed assessment of whether depressive disorder incidence contributed explanatory value beyond established population level determinants of dementia incidence. Use of standardised global datasets further supported broad cross-national comparability.

Several limitations should be acknowledged. As an ecological analysis, the findings cannot be interpreted at the individual level and may be subject to ecological fallacy. Cross national variation in diagnostic practices, healthcare access, and reporting may influence incidence estimates. This is particularly relevant for depressive disorders and may be more pronounced among males. Residual confounding by unmeasured cultural or health system factors may persist despite adjustment for major structural indicators. Moreover, the independent contribution of depressive disorder incidence in females was modest relative to dominant ageing-related determinants and should be interpreted as a supplementary population-level correlate rather than a primary driver of dementia burden.

### Implications for Sex-Stratified Epidemiological Analysis

The findings highlight the importance of explicit sex stratification in population-level dementia research. Treating sex solely as a covariate assumes equivalence in underlying pathways, whereas direct sex-comparative analysis can reveal meaningful divergence in how mental health conditions are embedded within structural and demographic contexts. The results suggest that depressive disorder incidence functions as a population-level vulnerability marker for dementia in females but not in males, underscoring the need for sex-sensitive interpretation of ecological associations.

More broadly, this study demonstrates the value of integrating multiple analytical approaches with structural adjustment when examining mental health–dementia relationships at the population level. Such frameworks can help distinguish shared structural co-variation from sex-specific patterns, supporting more nuanced interpretation of global dementia epidemiology and informing prevention and surveillance strategies that are sensitive to gendered life-course and structural contexts.

## Conclusion

This study demonstrates that the population-level association between depressive disorder incidence and dementia incidence differs substantially between females and males. Across multiple analytical approaches, depressive disorder incidence showed a consistently stronger, more structured, and more independent association with dementia incidence in females, whereas no comparable association was observed in males. These findings indicate that depressive disorder is not a uniform population-level correlate of dementia, but instead functions as a sex-specific risk indicator with primary relevance to female dementia burden.

By explicitly comparing sexes rather than treating sex as a covariate, this study clarifies how mental health conditions are differentially embedded within population-level dementia risk architectures. Female depressive disorder incidence appears to reflect broader population-level vulnerabilities that remain relevant to dementia incidence beyond shared ageing and structural determinants, while male depressive disorder incidence is largely decoupled from these pathways. Incorporating sex-stratified perspectives into global dementia epidemiology may therefore improve interpretation of cross-national dementia patterns and supports the need for dementia prevention and mental health surveillance frameworks that recognise depressive disorder incidence as a population-level indicator of dementia vulnerability among women, particularly in developing and structurally transitioning contexts.

## Data Availability

All data produced in the present study are available upon reasonable request to the authors.

https://ghdx.healthdata.org/record/global-burden-disease-study-2019-gbd-2019-covariates-1980-2019

## Ethics Statement

Ethical approval was obtained from the Office of Research Ethics, Compliance and Integrity (ORECI), University of Adelaide (Approval No. 36289).

## Consent statement

Not applicable due to aggregated data used for this study.

## Conflict of interest statement

The author declared that there is no conflict of interest.

## Author contribution statement

WY, as the sole author for this study reviewed the literature and obtained the data for structing this study. WY formulated the hypothesis relating nurse-midwifery density to neonatal mortality rate, analysed the data and interpreted results and wrote the text. WY approved the final version of the manuscript.

## Funding statement

This research did not receive any specific grant from funding agencies in the public, commercial, or not-for-profit sectors.

## Supplemental File

Table S1: Sex-specific correlations between depressive disorder incidence and dementia incidence across global, economic, and regional country groupings

## Notes

### Competing Interest Statement

The authors have declared no competing interest.

### Author Declarations

The data sources for this study are described in the Materials and Methods section. All data used are freely available from the official websites of international organizations. Formal permission to use the data for noncommercial research purposes was not required, as their use complies with the public access permissions outlined in the respective agencies terms and conditions.

