## Supplementary material for "Why Depression Matters More for Dementia in Females: Global Population Evidence": 2-2 Supplementary Table S1: 2-2 Supplementary Table S1.docx

Supplementary Table S1: Sex-specific correlations between depressive disorder incidence and dementia incidence across global, economic, and regional country groupings

| **Country groupings** | **Female:** depressive disorder incidence and dementia incidence | |  | **Male:** depressive disorder incidence and dementia incidence | | **n** |
| --- | --- | --- | --- | --- | --- | --- |
|  | **Pearson’s r** | **Spearman’s ρ** |  | **Pearson’s r** | **Spearman’s ρ** |  |
| Worldwide (All countries) | 0.340 (p < .001) | 0.341 (p < .001) |  | –0.053 (p = .450) | –0.061 (p = .387) | 204 |
| **World Bank income classifications** | | |  |  |  |  |
| High-income countries | **0.249** (p = .040) | **0.228** (p = .061) |  | **–0.112** (p = .364) | **–0.051** (p = .678) | 68 |
| Upper-middle-income countries | **0.344** (p = .011) | **0.314** (p = .021) |  | **–0.106** (p = .446) | **–0.096** (p = .488) | 54 |
| Lower-middle-income countries | **0.293** (p = .041) | **0.235** (p = .104) |  | **0.010** (p = .943) | **–0.054** (p = .714) | 49 |
| Low-income countries | **0.549** (p = .002) | **0.544** (p = .003) |  | **0.426** (p = .024) | **0.357** (p = .062) | 28 |
| Low- and middle-income countries (LMICs) | **0.299** (p < .001) | **0.316** (p < .001) |  | **–0.053** (p = .545) | **–0.073** (p = .410) | 131 |
| Fisher’s *r*-to-*z*: High-income vs LMICs | z= **–** 0.36 (p=0.7188) | z= **–** 0.62 (p=0.5353) |  | z=**–**0.39 (p= 0.3483) | z= 0.15 (p=0.8808) |  |
| **UN common practice** | |  |  |  |  |  |
| Developed countries | **0.244** (p = .092) | **0.279** (p = .053) |  | **0.172** (p = .237) | **0.197** (p = .176) | 49 |
| Developing countries | **0.306** (p < .001) | **0.330** (p < .001) |  | **–0.077** (p = .340) | **–0.057** (p = .479) | 155 |
| Fisher’s *r*-to-*z*: Developed vs developing | z=**–** 0.4 (p= 6892) | z=**–**1.72 (p=0.0854) |  | z= 1.19 (p=0.1362) | z= 0.85 (p=0.3953) |  |
| **WHO Regions** |  |  |  |  |  |  |
| African Region | **0.266** (p = .071) | **0.323** (p = .027) |  | **0.200** (p = .178) | **0.190** (p = .201) | 47 |
| Region of the Americas | **0.313** (p = .055) | **0.347** (p = .033) |  | **0.152** (p = .363) | **0.328** (p = .044) | 38 |
| Eastern Mediterranean Region | **0.516** (p = .017) | **0.578** (p = .006) |  | **0.359** (p = .110) | **0.418** (p = .059) | 21 |
| European Region | **0.138** (p = .323) | **0.171** (p = .220) |  | **0.155** (p = .266) | **0.150** (p = .285) | 53 |
| South-East Asia Region | **–0.300** (p = .370) | **–0.282** (p = .401) |  | **–0.143** (p = .675) | **0.118** (p = .729) | 11 |
| Western Pacific Region | **0.631** (p < .001) | **0.576** (p = .001) |  | **0.025** (p = .899) | **–0.049** (p = .802) | 29 |
| **Countries grouped based on various factors** | | |  |  |  |  |
| Asia Cooperation Dialogue | **–0.008** (p = .967) | **–0.149** (p = .440) |  | **–0.270** (p = .157) | **–0.300** (p = .114) | 29 |
| Asia-Pacific Economic Cooperation | **0.560** (p = .010) | **0.538** (p = .014) |  | **0.419** (p = .066) | **0.398** (p = .082) | 20 |
| Arab World | **0.588** (p = .005) | **0.705** (p < .001) |  | **0.476** (p = .029) | **0.577** (p = .006) | 21 |
| European Economic Are | **–0.006** (p = .975) | **0.003** (p = .986) |  | **0.313** (p = .098) | **0.272** (p = .154) | 29 |
| English, Official Language | **0.479** (p < .001) | **0.453** (p < .001) |  | **0.189** (p = .171) | **0.092** (p = .507) | 54 |
| European Union | **0.076** (p = .708) | **0.107** (p = .596) |  | **0.335** (p = .087) | **0.281** (p = .156) | 27 |
| Latin America | **0.402** (p = .051) | **0.383** (p = .065) |  | **0.112** (p = .603) | **0.249** (p = .241) | 24 |
| Latin America and the Caribbean | **0.308** (p = .072) | **0.338** (p = .047) |  | **0.098** (p = .575) | **0.305** (p = .074) | 35 |
| Organisation for Economic Co-operation and Development | **−0.071** (p = .674) | **−0.046** (p = .786) |  | **0.088** (p = .606) | **0.142** (p = .400) | 37 |
| Southern African Development Community | **0.027** (p = .921) | **0.091** (p = .737) |  | **0.011** (p = .968) | **0.100** (p = .713) | 16 |
| Shanghai Cooperation Organisation | **0.047** (p = .825) | **−0.101** (p = .632) |  | **0.006** (p = .978) | **−0.107** (p = .611) | 25 |

**Notes:** Pearson’s correlation coefficients (*r*) and Spearman’s rank correlation coefficients (ρ) quantify the sex-specific association between depressive disorder incidence and dementia incidence within each grouping. Pearson’s *r* reflects linear associations, while Spearman’s ρ captures monotonic relationships. Fisher’s *z* tests were used to compare correlation coefficients between groups.
